# A genome-wide investigation into the underlying genetic architecture of personality traits and overlap with psychopathology

**DOI:** 10.1101/2024.01.17.24301428

**Authors:** Priya Gupta, Marco Galimberti, Yue Liu, Sarah Beck, Aliza Wingo, Thomas Wingo, Keyrun Adhikari, VA Million Veteran Program, Murray B. Stein, Joel Gelernter, Daniel F. Levey

## Abstract

Personality is influenced by both genetic and environmental factors and is associated with other psychiatric traits such as anxiety and depression. The “Big Five” personality traits, which include neuroticism, extraversion, agreeableness, conscientiousness, and openness, are a widely accepted and influential framework for understanding and describing human personality. Of the big five personality traits, neuroticism has most often been the focus of genetic studies and is linked to various mental illnesses including depression, anxiety, and schizophrenia. Our knowledge of the genetic architecture of the other four personality traits is more limited. Utilizing the Million Veteran Program (MVP) cohort we conducted a genome-wide association study (GWAS) in individuals of European and African ancestry. Adding other published data, we performed GWAS meta-analysis for each of the five personality traits with sample sizes ranging from 237,390 to 682,688. We identified 158, 14, 3, 2, and 7 independent genome-wide significant (GWS) loci associated with neuroticism, extraversion, agreeableness, conscientiousness, and openness, respectively. These findings represent 55 novel loci for neuroticism, as well as the first GWS loci discovered for extraversion and agreeableness. Gene-based association testing revealed 254 genes showing significant association with at least one of the five personality traits. Transcriptome-wide and proteome-wide analysis identified altered expression of genes and proteins such as *CRHR1, SLC12A5, MAPT*, and *STX4*. Pathway enrichment and drug perturbation analyses identified complex biology underlying human personality traits. We also studied the inter-relationship of personality traits with 1,437 other traits in a phenome-wide genetic correlation analysis, identifying new associations. Mendelian randomization showed positive bidirectional effects between neuroticism and depression and anxiety while a negative bidirectional effect was observed for agreeableness and these psychiatric traits. This study improves our comprehensive understanding of the genetic architecture underlying personality traits and their relationship to other complex human traits.

## INTRODUCTION

Personality dimensions influence behavior, thoughts, feelings and reactions to different situations. A valuable construct within the field of psychological research has converged on five different dimensions to characterize human personality: neuroticism, extraversion, agreeableness, conscientiousness and openness [1, 2]. Personality dimensions could be playing an important role in the susceptibility and resilience to diagnosis of psychiatric disorders and their relationship with other health-related traits and responses to treatment.

The last decade has seen increasing interest in understanding the dimensions of human personality through the lens of genetics. Depression is one mental disorder that has been studied with respect to its relationship to personality traits, with a large portion of genetic risk for depression being captured by neuroticism [3]. The same study found a modest negative association of genetic depression risk with conscientiousness, with small contributions from openness, agreeableness, and extraversion. Neuroticism is one of the most studied dimensions of the “Big Five” and numerous studies have found positive correlations with depression, anxiety, and other mental illnesses [3–5]. Schizophrenia has also been associated with personality traits, especially neuroticism, which has been shown to increase risk for diagnosis [6]. A study using data from the Psychiatric Genomics Consortium (PGC) and personal genomics company 23andMe found two genomic loci to be common between neuroticism and schizophrenia. This study also reported six loci shared between schizophrenia and openness [7].

The past 15 years have seen an explosion in the use of the genome-wide association study (GWAS). In 2010, Marleen de moor *et al.* from the Genetics of Personality Consortium (GPC) published a GWAS study of Big Five personality traits conducted with 17,375 adults from 15 different samples of European ancestry (EUR) [8]. This study found two genome-wide significant (GWS) significant variants near the *RASA1* gene on 5q14.3 for openness and one near *KATNAL2* on 18q21.1 for conscientiousness but no significant associations for other personality traits. GPC then conducted studies on extraversion and neuroticism in their second phase and meta-analyses were performed. A GWAS of neuroticism that was conducted on approximately 73,000 subjects identified rs35855737 in the *MAG1* gene as a GWS variant [9]. Though the sample size was increased significantly to 63,030 subjects in phase-II, no GWS variants were detected for extraversion [10]. The largest GWAS study available to date for any personality trait is the neuroticism meta-analysis study published by Nagel *et al*. in 2018 [11]. In that study, the authors collected neuroticism genotype data of 372,903 individuals from the UK Biobank (UKB) and a meta-analysis was performed by combining the summary statistics from this UKB sample, 23andMe and GPC-phase 1 samples, increasing the total sample size to 449,484. They identified a total of 136 loci and 599 genes showing GWS associations to neuroticism.

In this work, we conducted GWAS of each of the Big Five personality traits in a sample of ∼224,000 individuals with genotype data available from the MVP. Using linkage disequilibrium score regression (LDSC), we estimated the SNP-based heritability of each of the 5 personality traits. We then combined the MVP data with other sources of personality GWAS summary statistics from GPC and UKB and performed meta-analyses for each of the five personality traits, including as many as ∼680,000 participants for the largest meta-analysis to date of neuroticism.

To gain insights into the biology of these traits, we performed transcriptome-wide association studies (TWAS) and proteome-wide association studies (PWAS) followed by pathway and drug perturbation analyses and variant fine mapping. We also studied the overlap of these personality traits with anxiety and other complex traits through phenome-wide genetic correlations and conditional analyses. We performed drug perturbation analyses with genes associated with neuroticism and found convergence on drugs for MDD. Finally, we conducted Mendelian randomization (MR) experiments to investigate the causal relationship of neuroticism and agreeableness, the two most genetically divergent traits, with depression and anxiety.

## RESULTS

### MVP GWAS

In the EUR GWAS in the MVP cohort, we identified in total 30 unique independent genomic loci significantly associated (P value < 5 × 10-8) with at least one of the five personality traits (see Table 1). The highest numbers of loci were found for extraversion and neuroticism (11 and 7 respectively) while conscientiousness showed only 2 loci. In the MVP we identified 4036 GWS variants (P<5 × 10^-8^) for neuroticism across 7 independent genomic loci harboring genes including *MAD1L1, MAP3K14, CRHR1, CRHR1-IT1,* and *VK2* (P<5X10^-08^). We identified 11 GWS loci for extraversion, the first GWS loci to be identified for this trait, to our knowledge.

**Table 1:** Genomic loci identified in the MVP cohort for the five personality traits.

|  | Genomic Locus | uniqID (CHR:Position) | P-value | Start | End | LeadSNPs | Genes |
| --- | --- | --- | --- | --- | --- | --- | --- |
| Neuroticism |  |  |  |  |  |  |  |
|  | 1 | 2:58156539:C:T | 7.87E-10 | 57942987 | 58484172 | rs2139053 | VRK2;BCL11A;FANCL |
|  | 2 | 7:2076701:C:T | 3.15E-10 | 1932629 | 2110850 | rs6948912 | FTSJ2;MAD1L1;AC110781.3 |
|  | 3 | 8:10903475:A:T | 4.39E-12 | 8088230 | 11895484 | rs7825636;rs615632;rs2001433 | TDH;FAM167A;MSRA<br>TDH;FAM167A;MSRA |
|  | 4 | 8:143312933:A:C | 1.80E-15 | 1.43E+08 | 1.44E+08 | rs4129585;rs117713019 | NA |
|  | 5 | 11:13328979:C:G | 1.17E-11 | 13230633 | 13350131 | rs7396943 | ARNTL |
|  | 6 | 16:61833811:C:T | 3.24E-08 | 61775810 | 61953413 | rs6498809 | CDH8 |
|  | 7 | 17:43667635:A:G | 3.95E-14 | 43460181 | 44865603 | rs574307253;rs116956554 | KANSL1;CRHR1;MAPT |
| Extraversion |  |  |  |  |  |  |  |
|  | 1 | 1:71834574:G:T | 1.13E-08 | 71822831 | 71896762 | rs11209774 | NA |
|  | 2 | 2:58167698:G:GA | 1.11E-08 | 57942987 | 58478157 | rs5831479 | VRK2;BCL11A |
|  | 3 | 2:185130889:A:G | 1.79E-08 | 1.85E+08 | 1.85E+08 | rs7606514 | NA |
|  | 4 | 4:8536348<br>9:C:T | 4.8<br>1E-08 | 8532<br>6010 | 8540<br>8000 | rs1444978 | NA |
|  | 5 | 5:9300872<br>2:C:T | 5.6<br>1E-09 | 9297<br>7139 | 9342<br>7260 | rs1011501 | FAM172A |
|  | 6 | 6:9231531<br>7:G:T | 4.5<br>5E-08 | 9223<br>4049 | 9234<br>0527 | rs7739331 | NA |
|  | 7 | 11:132487<br>30:C:T | 5.6<br>4E-10 | 1323<br>0633 | 1334<br>5926 | rs35424804 | ARNTL |
|  | 8 | 12:108618<br>630:C:T | 4.9<br>7E-09 | 1.09<br>E+08 | 1.09<br>E+08 | rs3764002 | WSCD2 |
|  | 9 | 17:437786<br>80:A:T | 2.4<br>2E-09 | 4346<br>3493 | 4477<br>3783 | rs17688916 | KANSL1;CRHR1;M<br>APT |
|  | 10 | 17:794527<br>56:A:AT | 7.9<br>4E-09 | 7942<br>9358 | 7947<br>3743 | rs35918640 | FSCN2;RP13-766D20.2 |
|  | 11 | 19:318766<br>92:A:C | 4.3<br>3E-09 | 3183<br>0613 | 3188<br>5009 | rs12971383 | TSHZ3;AC007796.1 |
| Agreeable<br>ness |  |  |  |  |  |  |  |
|  | 1 | 7:1142108<br>14:C:T | 4.0<br>0E-10 | 1.14<br>E+08 | 1.14<br>E+08 | rs17137124 | FOXP2;MIR3666 |
|  | 2 | 8:1070026<br>6:G:T | 4.0<br>6E-09 | 1051<br>6336 | 1145<br>0422 | rs7833945 | TDH;FAM167A;SOX<br>7 |
|  | 3 | 18:531952<br>49:A:G | 1.9<br>2E-08 | 5319<br>5249 | 5346<br>3661 | rs7240986 | RP11-397A16.1 |
| Conscient<br>iousness |  |  |  |  |  |  |  |
|  | 1 | 7:1142971<br>80:A:G | 1.5<br>1E-08 | 1.14<br>E+08 | 1.14<br>E+08 | rs936145 | FOXP2;MIR3666 |
|  | 2 | 8:8144346<br>1:A:G | 8.6<br>2E-09 | 8135<br>8464 | 8144<br>3461 | rs78446248 | NA |
| Openness |  |  |  |  |  |  |  |
|  | 1 | 2:2937792<br>3:A:T | 2.0<br>3E-08 | 2934<br>5180 | 2940<br>3624 | rs6725323 | NA |
|  | 2 | 4:1529456<br>67:C:T | 2.7<br>7E-09 | 1.53<br>E+08 | 1.53<br>E+08 | rs919013 | NA |
|  | 3 | 6:1359287<br>72:G:T | 4.2<br>9E-10 | 1.36<br>E+08 | 1.36<br>E+08 | rs117890891 | AHI1 |
|  | 4 | 8:6546344<br>2:C:T | 3.4<br>4E-09 | 6543<br>7964 | 6549<br>8165 | rs6996198 | ARMC1;RP11-21C4.5;CYP7B1 |
|  | 5 | 8:1416472<br>91:A:C | 3.9<br>2E-08 | 1.42<br>E+08 | 1.42<br>E+08 | rs11996715 | PTK2;TRAPPC9;RP1<br>1-809O17.1 |
|  | 6 | 9:3577744<br>2:G:GTT | 2.5<br>6E-08 | 3573<br>1560 | 3585<br>1048 | rs61689447 | TMEM8B;NPR2;TP<br>M2;GBA2;CREB3<br>;SPAG8 |
|  | 7 | 11:666106<br>45:C:G | 2.5<br>4E-13 | 6606<br>6349 | 6667<br>3079 | rs7570 | PC;LRFN4;C11orf80 |

Associations for extraversion were found near several genes including *CRHR1, MAPT*, and *METTL15* (total 90 genes). For the 2 conscientiousousness, loci,the first locus maps to a region near the genes *FOXP2, PPP1R3A*, and *MDFIC* and the second locus maps to *ZNF704* gene – all are protein coding genes. For openness, 7 loci were identified spanning over 39 genes including *BRMS1, RIN1*, and *B3GNT1*. For agreeableness, 3 loci were identified spanning 19 genes including *SOX7, PINX1,* and *FOXP2.*The Manhattan plots for all 5 traits are shown in Supplementary Fig. S1.

One GWS variant was found for agreeableness in the African Ancestry (AFR) sample. This variant rs112726823 (P<3.27E-08) mapped near *ARHGAP24*. We did not find any GWS variants for any of the other four personality traits in the AFR the multiple subthreshold findings from this analysis may reach the GWS threshold in a larger sample. A list of top scoring independent SNPs found in the AFR sample for each trait is provided in Supplementary Sheet 1.

### Meta-analysis in European ancestry populations

The meta-analysis for neuroticism showed associations with 158 independent GWS loci. The increased power due to the inclusion of MVP data resulted in the identification of 55 additional GWS novel loci which were not significant in the previous study [11]. Only 3 loci identified previously (rs1763839, rs2295094, rs11184985) were no longer significant in our meta-analysis. SNPs and loci were mapped to genes based on chromosomal position, eQTL and chromatic interaction information using FUMA [12]. 231 genes were found significant in the MAGMA gene-based test [13]. *NSF, KANSL1, CRHR1, FMNL1*, and *PLEKHM1* (P<2.85 × 10^-40^) were among the top significant hits. The largest number of significant loci are located on chromosome 17, followed by chromosome 18.

For extraversion, after meta-analyzing the MVP and UKB data, the number of significant loci increased to 10. The lead signals were located on chromosomes 1, 2, 5, 11, and 17. The most significant locus harbors genes in/near *WSCD2* (P < 3.45 × 10^-11^) located on chromosome 12.

Chromosome 11 contains significant variant associations from three traits-neuroticism, extraversion and agreeableness, with neuroticism and extraversion both having findings near the “basic helix-loop-helix ARNT like 1” (*ARNTL1*, also known as *BMAL1*) gene, with opposing and significant direction of effect at common variants. Complete information of all identified significant loci for each of the five traits is provided in Supplementary Sheet S2.

### Linkage Disequilibrium Score Regression

We first used LDSC to calculate SNP-based heritability of each of the five personality traits within the MVP EUR cohort. The SNP-heritability ranges from 4% to 7% (see Supplementary Fig. S2), with extraversion showing the highest heritability point estimate of all traits (neuroticism h2=6%, agreeableness h2=4%, extraversion h2=7%, openness h2=5%, conscientiousness h2=5%). The values were not significant for AFR, likely owing to its lower power due to its small sample size.

Before combining the MVP cohort-derived summary statistics with other data sources, we calculated the genetic correlation between the MVP personality summary statistics and other respective sources (see Table S1). A correlation coefficient value of 0.80 observed for the neuroticism summary statistics from the MVP cohort and Nagel *et al.* study[11] suggests that there is limited heterogeneity between the two datasets and supports their use in a meta-analysis. As shown in Supplementary Table S1, the genetic correlations were high for all other 4 traits across data sources as well.

LDSC was used to estimate SNP-based heritability in the EUR participants for each personality trait in the meta-analysis. The SNP-heritability values in the meta analyses were similar to what was observed in the MVP-only cohort for the different traits in the EUR with a decrease in heritability of extraversion from 7.1% to 5.1% (see Fig 1B).

**Figure 1:**
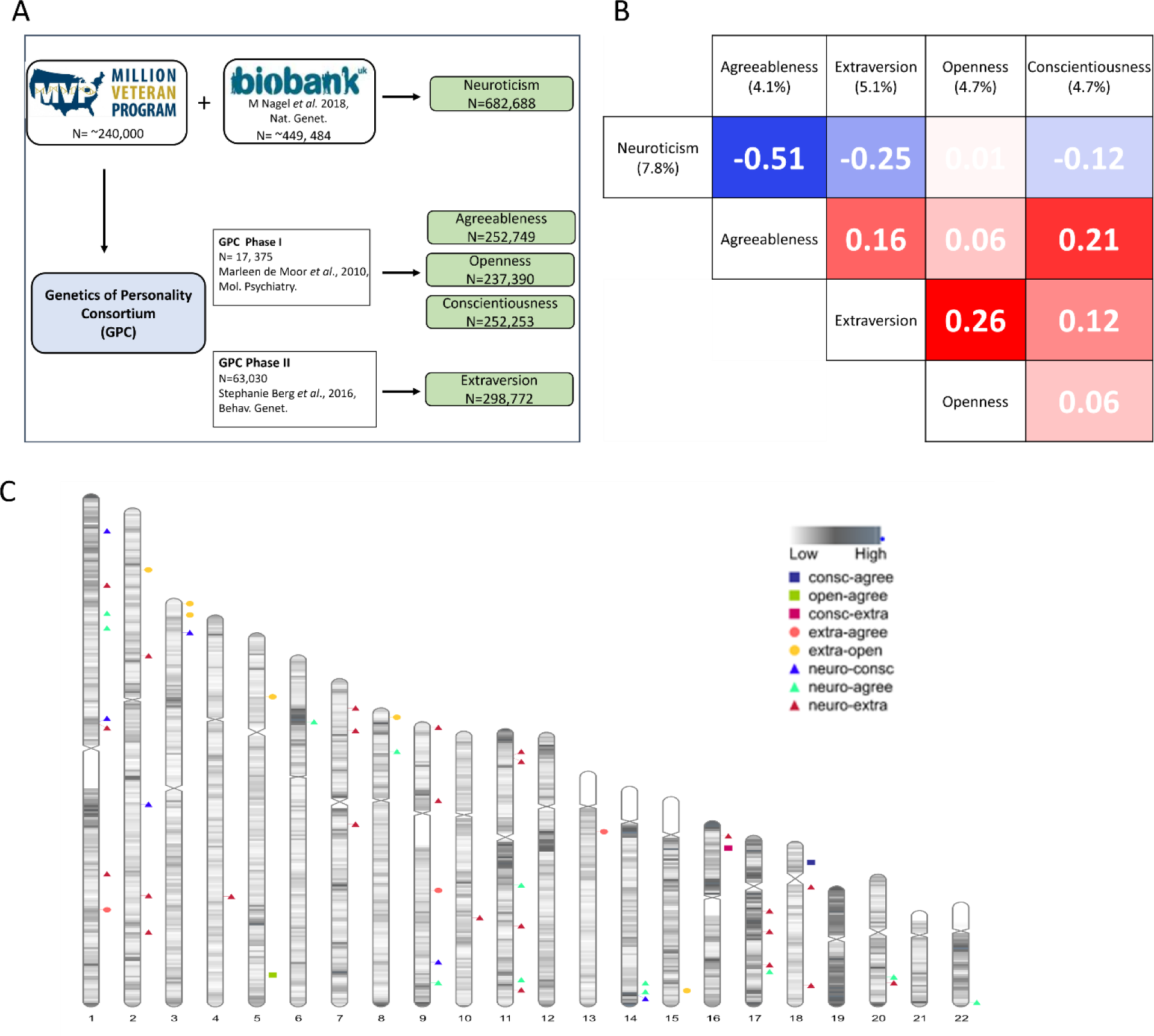
(A) Data collection of the 5-personality traits (B) Genetic correlation matrix among the 5 personality traits (metadata). The heritability value of the respective trait is written in parenthesis. (C) Karyogram showing the regions with significant local genetic correlation (rG>0.3) between different personality traits.

**Figure 2:**
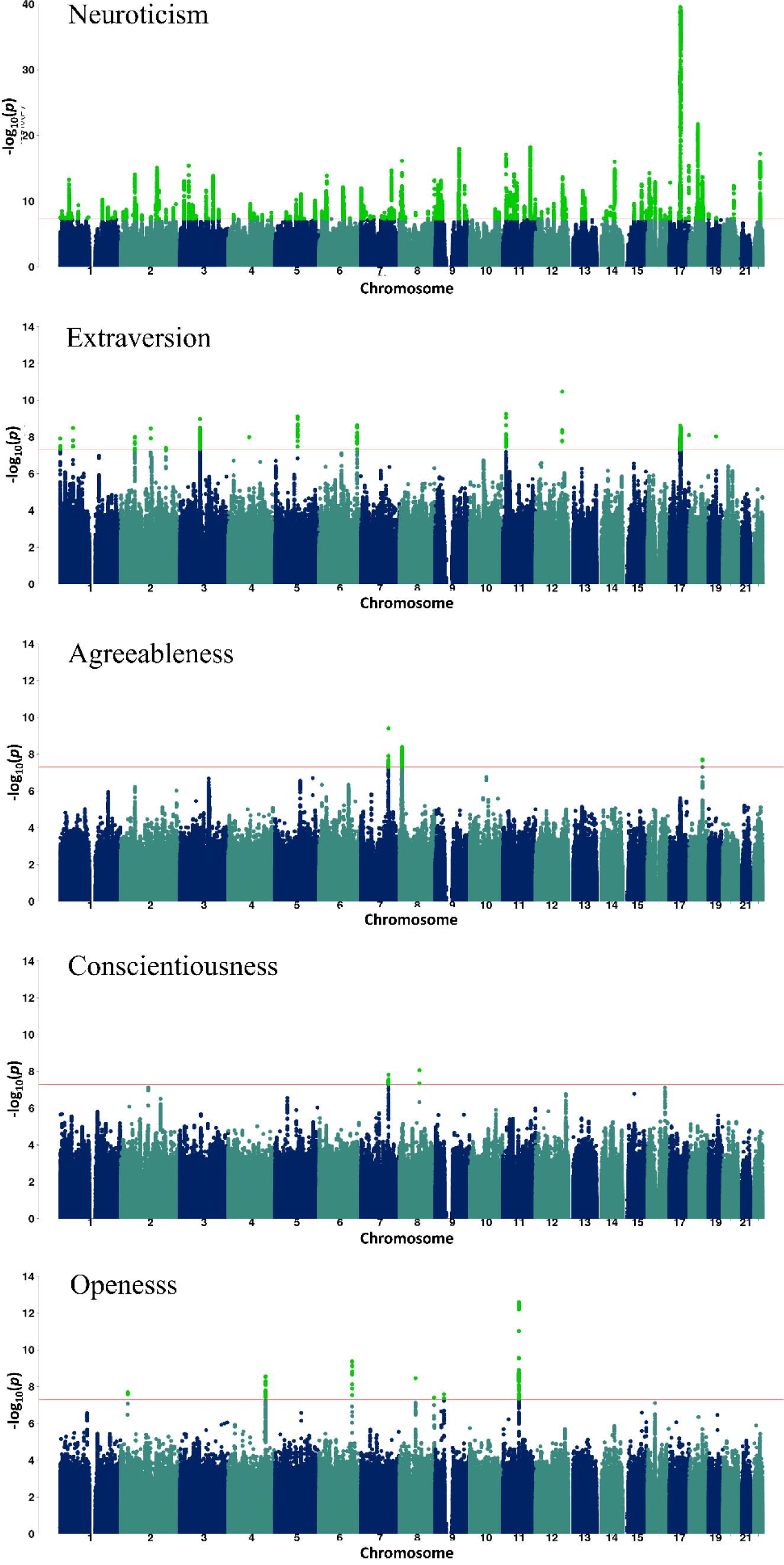
Metaanalysis data GWAS Manhattan plots of the 5 personality traits showing the genome-wide significant (GWS) variants in light green color. The red line depicts the p-value threshold used.

Genetic correlation estimates were also obtained between the meta-analysis summary statistics for the 5 personality traits . We found a significant degree of varying genetic overlap among the 5 personality traits. The genetic correlations are presented in Fig 1B. The highest correlation is observed between neuroticism and extraversion with a rG= -0.51.

Next, we estimated the genetic correlations of 1,437 traits listed in the Complex Traits Genetics Virtual Lab (CTG-VL)[14] summary statistics record to find other traits related to the 5 personality traits (see Supplementary Sheet 3). Three hundred twenty-five traits showed significant genetic correlation to one or more personality traits. We found MDD and anxiety showed varying degree of significant correlations to different personality traits as shown in Fig 5. The highest genetic correlation is between neuroticism and anxiety (rG=0.80). Neuroticism and agreeableness both show high genetic correlations to these traits but in opposite directions: MDD (Neuroticism rG=0.68, Agreeableness rG=-0.35), manic behavior (Neuroticism rG=0.44, Agreeableness rG=-0.35), anxiety (Neuroticism rG=080, Agreeableness rG=-0.33), and irritability (Neuroticism rG=0.70, Agreeableness rG=-0.62).

### Local genetic correlations

Global genetic correlations use the average squared signal over the entire genome which may sometimes mask opposing local correlations in different genomic regions. To counter that, we also calculated the local genetic correlations among the 5 personality trait pairs using LAVA [15]. All personality pairs showed varying degree of correlation in different genomic regions except for the neuroticism-openness pair, which showed negligible global (rG=-0.01) and no local genetic correlation between the two. The highest number of correlated genomic chunks were found for neuroticism-extraversion and neuroticism-openness pairs (see Fig. 1C & Supplementary Sheet 4).

### TWAS

We performed TWAS for each of the Big Five personality traits in EUR (meta-analysis) using FUSION [16] and the GWAS summary statistics. We performed a multi-tissue TWAS in 13- different brain sub-tissues and blood using their respective expression profiles from GTEx v8 [17]. From a total 10,890 genes tested, we identified total 175, 24, 5, 1, 11 genes, respectively, showing significant gene-trait associations across the 13 sub-tissues in neuroticism, extraversion, agreeableness, conscientiousness, and openness after Bonferroni correction for 141,570 tests (10,890 genes across 13 tissues) (see Fig 3A). Figure 3A shows the distribution of associations found across the 13 tissues for each trait. The highest number of gene-trait associations were found in brain caudate basal ganglia, cerebellum, cerebral hemisphere and frontal cortex regions for neuroticism and extraversion while fewer TWAS gene-trait associations were identified for the other three personality traits, presumably owing to the comparatively lower power of their respective GWAS datasets.

**Figure 3:**
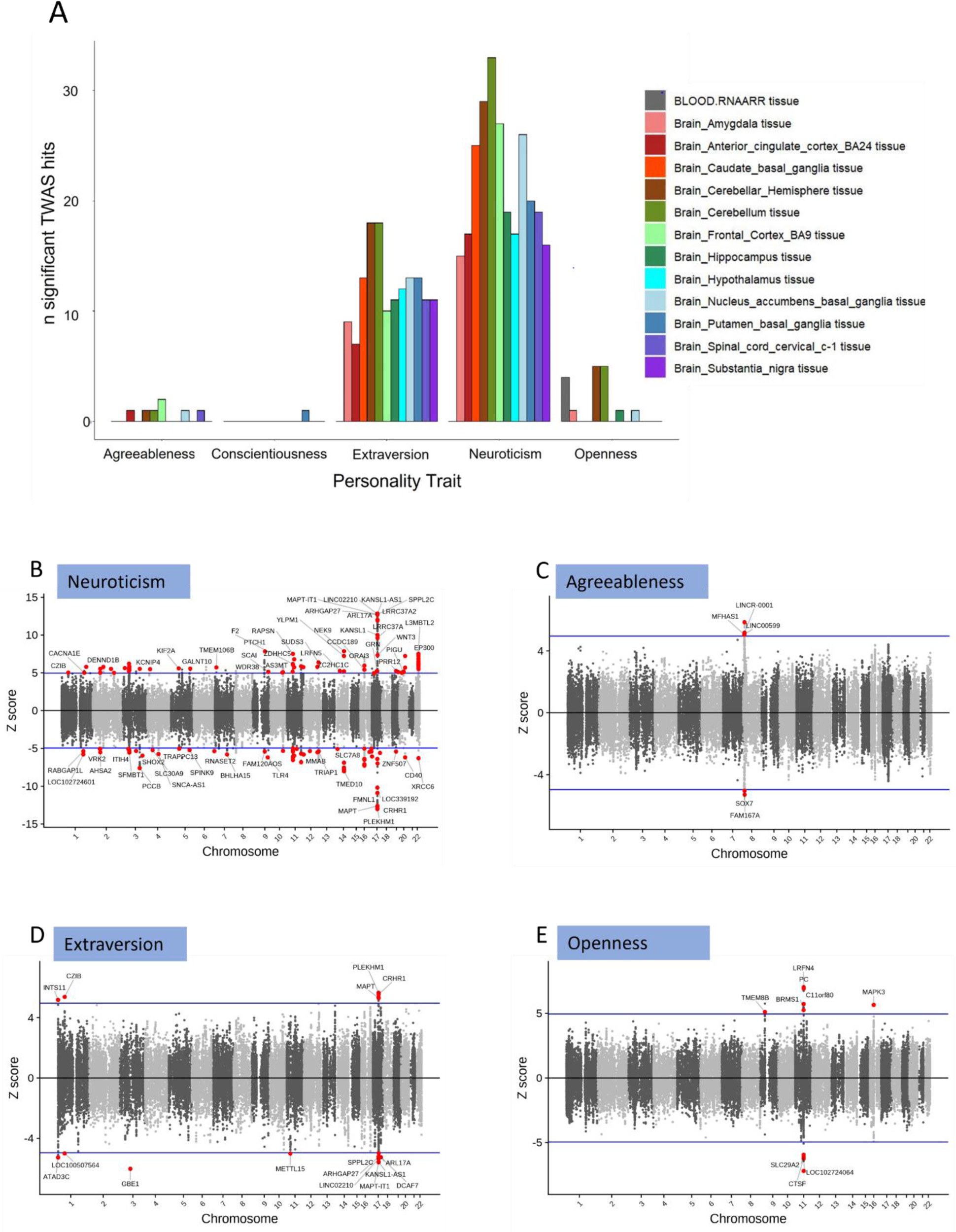
(A) bar chart showing the number of significant TWAS genes/transcripts found four personality traits with significant findings in respective sub-tissues. (B-E) scatter plots with TWAS z-scores of each gene/transcript plotted on y-axis and its respective chromosomal location plotted on x-axis. The significant hits are shown in red circles with mapped gene names as labels. The blue horizontal line indicates the significance threshold of z-score corresponding to Bonferroni-corrected p-value. Conscientiousness data is reported in Supplemental Sheet 4.

*CRHR1, KANSLI1-AS1, MAP-IT1* are among the top TWAS gene associations (P<3.69X10^-08^) for neuroticism (Figure 3B). The strong association of *CRHR1* (encoding corticotropic releasing hormone receptor) which in some prior work has been shown to be associated with treatment response to depression may suggest[18] some common underlying elements regulating both neuroticism and depression. Extraversion also shows strong gene-trait associations with *CRHR1, KANSL1-AS1 and MAPT-IT1* but with an opposite direction of effect to neuroticism. This may indicate some common genetic components whose differential behavior regulates neuroticism and extraversion. There are nine such genes showing opposite direction of effect in neuroticism and extraversion.

*LOC10271024064* and *LRFN4* showed the strongest associations with openness and *LINCR-0001* and *FAM167A* showed the strong associations with agreeableness while only one gene- *AP1G1* showed association with conscientiousness in the 13 tissues considered. The complete list of all GWS TWAS gene hits for the five personality traits is provided in Supplementary Sheet 5.

### PWAS

We investigated the association of personality traits with protein expression using PWAS. Based on the availability of protein profiles and the observed TWAS signal, dorsolateral prefrontal cortex (dlPFC) brain protein profiles were chosen for the PWAS analysis. The PWAS identified 47 proteins to be significantly associated with neuroticism. Next, we checked the colocalization signal for these PWAS lead genes. Out of 47 PWAS lead genes, 35 genes showed a colocalization signal (HH4>0.5).

Five, two, two and four proteins were discovered for extraversion, agreeableness, conscientiousness, and openness, respectively (Fig 4). A complete list of all PWAS lead genes is provided in Supplementary Sheet 6.

**Figure 4:**
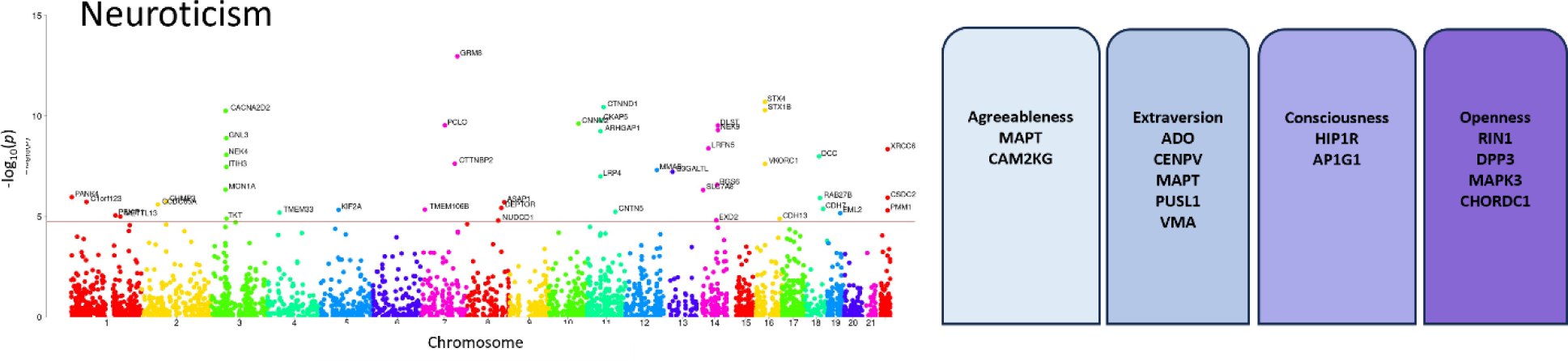
PWAS analysis. A Manhattan plots is displayed showing the significant protein associations observed for neuroticism. The boxes on the right shows the significant proteins found for the resp. 4 personality traits.

### Variant Fine mapping

To identify well-supported possible causal variants from the large list of SNPs showing associations with the personality traits, we performed genome-wide variant fine-mapping using PolyFun [19]. In total, 166 unique variants were fine mapped across the 5 personality traits. The number of variants fine mapped for neuroticism, extraversion, agreeableness, conscientiousness, openness were 155, 8, 4, 7, and 3, respectively. The complete list of variants fine-mapped for each of the personality trait is provided in the Supplementary Sheet 7.

### Relationship between personality traits and psychiatric disorders

We performed additional analyses to help understand the significant differential genetic correlation observed between neuroticism and agreeableness with different psychiatric-disorders like major depressive disorder (MDD) and anxiety.

### Conditional Analysis

Because the genetic correlation between anxiety and neuroticism was so high, we performed multi-trait-based conditional and joint analysis (mtCoJo) of neuroticism summary statistics conditioned on anxiety and MDD summary statistics individually. The Anxiety and MDD summary statistic used is based on data from UKB, MVP, PGS with individuals of EUR ancestry (see Methods for details). We performed a similar analysis with agreeableness, which had a negative correlation with both MDD and anxiety, as a negative control.

After conditioning on MDD, the SNP-heritability of the conditioned neuroticism summary statistic reduced significantly from 7.8% to 3%. Out of the original 158 GWS leads, only 37 remained significant after conditioning indicating there is substantial genetic overlap between neuroticism and MDD which gets removed after conditioning. In case of conditioning on anxiety, again there is a decrease in neuroticism heritability, but to a lesser extent (see Table 2). On conditioning agreeableness on MDD and anxiety, no significant reduction in heritability was observed. However, loss of one genomic locus – rs7240986 (18:53195249:A:G) was observed after conditioning on either anxiety or MDD for agreeableness.

**Table 2:** Conditional analysis.

| Primary Trait | Trait conditioned on | h2 (se) | No. of GWS loci before conditioning | h2 (se) after conditioning | Z-diff | P. value | No. of GWS loci after conditioning |
| --- | --- | --- | --- | --- | --- | --- | --- |
| Neuro | Anxiety | 0.078<br>(0.003) | 159 | 0.05 (0.001) | 8.85* | 8.41e-19 | 79 |
| Agree | Anxiety | 0.041<br>(0.003) | 3 | 0.034<br>(0.003) | 1.65 | 0.01 | 2 |
| Neuro | MDD | 0.078<br>(0.003) | 159 | 0.03 (0.002) | 13.31* | 1.95e-40 | 37 |
| Agree | MDD | 0.041<br>(0.003) | 3 | 0.036<br>(0.003) | 1.18 | 0.24 | 2 |

### Drug Perturbation Analysis

We performed a drug perturbation analysis to find drug candidates for neuroticism-enriched genes using gene2drug software [20]. Gene2drug utilizes the Cmap transcriptomics data of ∼13,000 cell lines exposed to different drugs and based on these gene expression profiles and then pathway expression profiles (PEPs), it first matches the query gene to its pathway and then to its potential candidate drug. This analysis predicted 298 unique drugs to correspond to the 231 significantly associated neuroticism genes. The top-scoring drug was found to be desipramine which is a tricylic antidepressant. Some of the other drugs predicted are flupenthixol (antipsychotic), tetryzoline (alpha-adrenergic agonist), doxorubicin (anthracycline/chemotherapy), and digitoxigenin (cardenolide). Based on these results, we repeated the drug perturbation analysis with depression enriched genes. While there were only 51 genes common between neuroticism and depression gene sets there was a convergence on drugs in the perturbation analysis. Out of 286 and 298 drugs predicted for depression and neuroticism, respectively, 167 drugs were common to both. The complete list of drugs is presented in Supplementary Sheet 8.

### Mendelian Randomization (MR)

After establishing genetic overlap of neuroticism with MDD and anxiety, we carried out an MR analysis to explore the possibility of a causal relationship between genetic risk for neuroticism and MDD or anxiety. The results of the MR analysis using different methods are presented in table 3A. The results of MR indicate a bidirectional causal effect, with the exposure of MDD on neuroticism outcome showing an IVW effect value of 0.429 at a significant P-value (2.07e-85). The exposure of neuroticism on MDD shows a higher causal effect value of 0.834 with a significant P-value (6.41e-103). We performed sensitivity analysis of MR using MRlap, which corrects for different sources of bias including sample overlap, because there is overlapping participants between the exposure and outcome datasets [21]. With MRlap, we observe similar results with positive significant corrected beta values in MRlap performed between MDD and neuroticism in both directions (see Table 3B).

**Table 3A:**
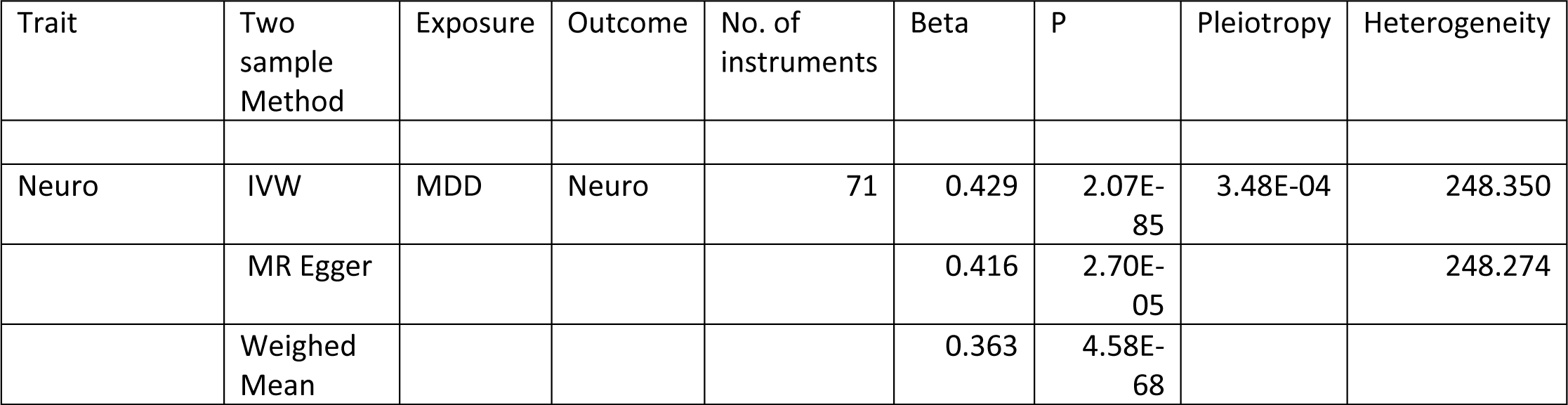
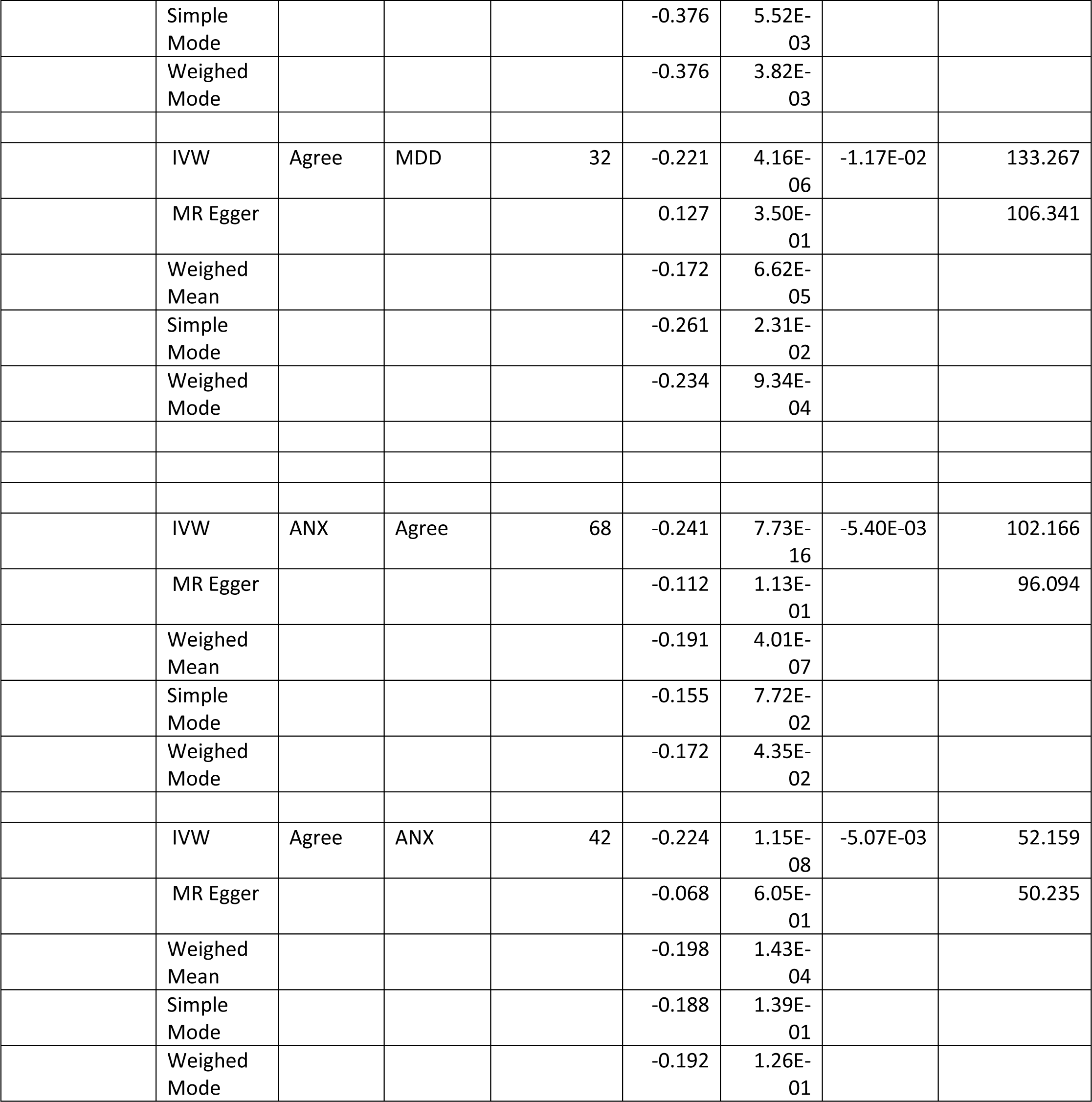
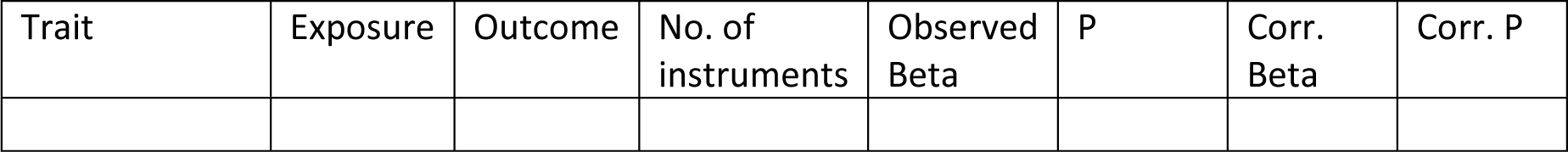
Outcome of mendelian randomization experiments performed using MR.

**Table 3B:**
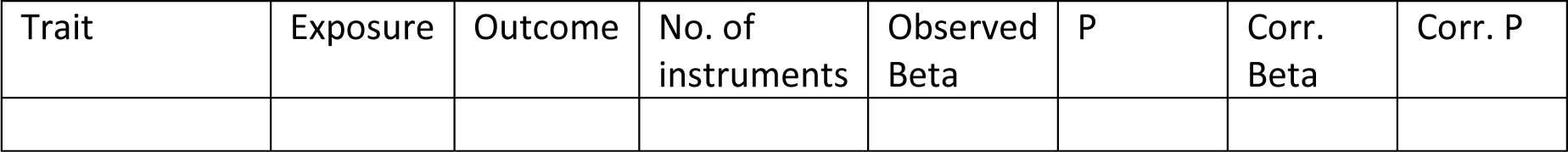
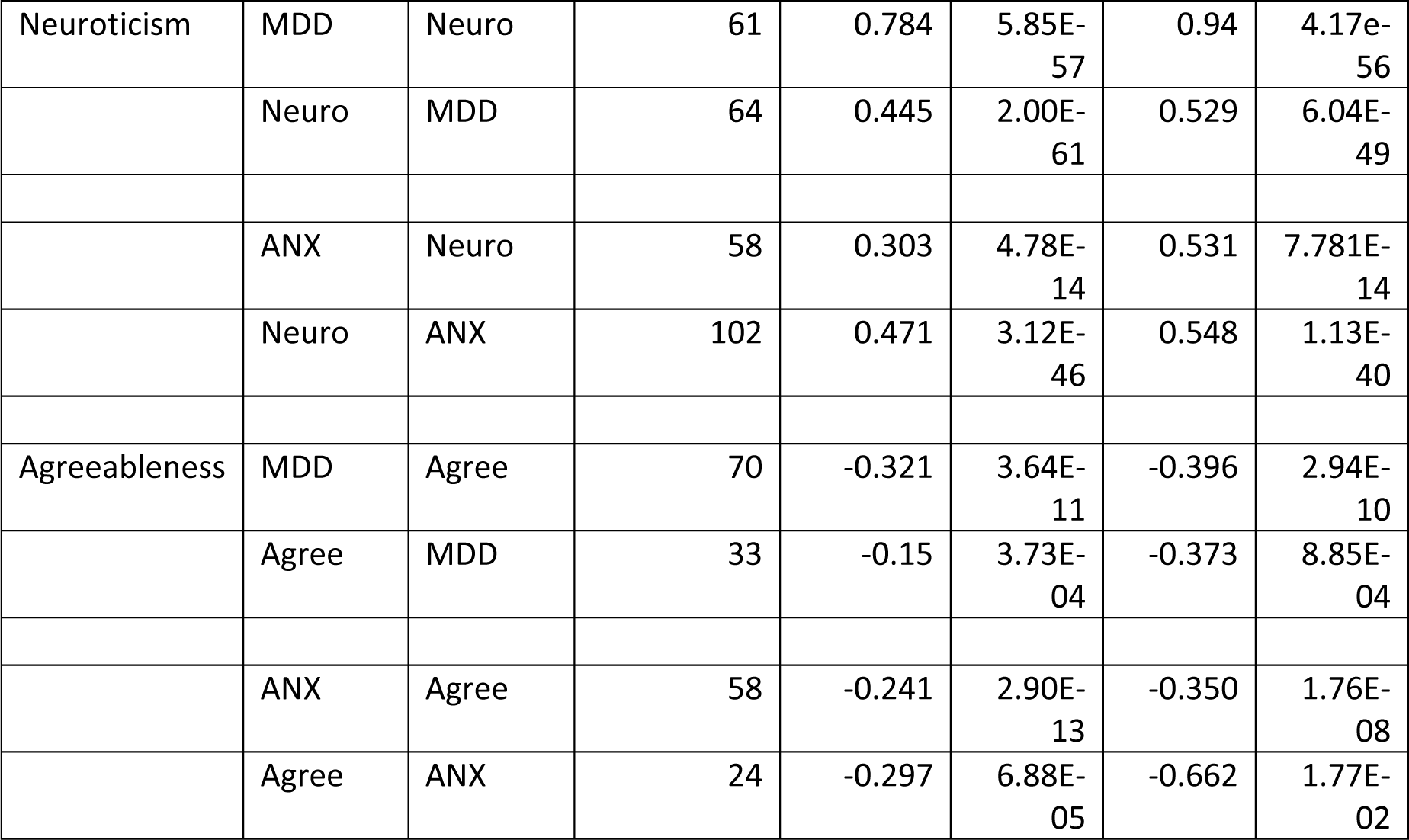
Outcome of MRLap sensitivity analysis.

We also investigated the casual relationship of neuroticism with anxiety. On performing MR with anxiety exposure on neuroticism, we found a beta value of 0.179 (p=1.25e-15) and a corrected beta value with MRlap of 0.531 (p=7.78e-14) showing evidence of causality. On reversing the direction, the causality effect was stronger as seen by higher beta value of 0.70 (p=5.76e-61) with MR and corrected beta value of 0.548 (p=1.13e-40) with MRlap. This suggests that there is stronger evidence of causal effect of neuroticism on anxiety as compared to the reverse based on the genetic susceptibility. GWAS studies of anxiety and anxiety disorders are still relatively underpowered compared to neuroticism, limiting the number of available genetic instruments available for testing as exposures.

We investigated the causal effect of agreeableness on MDD and anxiety and vice-versa. In case of MR of MDD exposure on agreeableness outcome, a beta value of -0.284 (p=5.77e-13) was observed indicating negative causal effect of MDD on agreeableness (table 3A and 3B). The causal effect is bidirectional with similar values observed in the opposite direction as well. The results are consistent with genetic correlation findings where negative correlation was observed between agreeableness and MDD. MR analysis of agreeableness and anxiety also indicated bidirectional causal effect. However, here both the traits have limited instruments available.

## METHODS

### Cohort and Phenotype

We used data release version 4 of the MVP [22]. The 10-item Big Five Inventory (BFI-10) was included as part of a self-report Lifestyle survey provided to MVP participants, with 2-items for each of the personality traits [23](**Supplementary Table S3**).

### Genotyping and Imputation

Genotyping and imputation of MVP subjects has been described previously [22, 24]. Briefly, a customized Affymetrix Axiom Array was used for genotyping. MVP genotype data for biallelic SNPs were imputed using Minimac4 [25] and a reference panel from the African Genome Resources (AGR) panel by the Sanger Institute. Indels and complex variants were imputed independently using the 1000 Genomes (1KG) phase 3 panel [26] and merged in an approach similar to that employed by the UK Biobank. Designation of broad ancestries was based on genetic assignment with comparison to 1KG reference panels [26].

### GWAS and Meta-Analysis

We performed individual GWAS for each of the five personality traits in the MVP cohort [22]. The personality information along with genotype data was available for 265,000 individuals, 235,000 EUR and 30,000 AFR. The GWAS was performed separately for each of the traits in the EUR and AFR datasets and the effect values were computed using linear mixed modelling algorithm.

MVP GWAS was conducted using linear regression in PLINK 2.0 using the first 10 PCs, sex, and age as covariates [27]. Variants were excluded if call missingness in the best-guess genotype exceeded 20%. Alleles with minor allele frequency (MAF) <0.1% were excluded. After applying all filters, genotype data from 233,204, 235,742, 235,374, 234,880 and 220,015 participants were included for neuroticism, extraversion, agreeableness, conscientiousness, and openness, respectively.

For meta-analysis, summary statistics generated in this study (referred as MVP study) were combined using METAL [28] with that from UKB and GPC phase-I and-II studies (see Figure 1A) based on the availability of data for respective traits. Inverse-variance weighing scheme of METAL was applied to weight the effect sizes of SNPs from different source studies. For neuroticism, summary statistics from MVP and UKBiobank study [11] were combined, increasing the total sample size to 682,688. For extraversion, summary statistics from MVP and GPC phase-II study [10] were combined while summary statistics from MVP and GPC phase-I study [8] were combined for the respective meta-analysis of agreeableness, openness and conscientiousness. There is a sample overlap between neuroticism GWAS summary statistics of GPC and the UKBiobank study, thus GPC derived summary statistics was not included in our meta-analysis of neuroticism.

### LDSC and SNP-Heritability

Linkage Disequilibrium Score Regression (LDSC) was performed based on the LD reference from the 1000 genome data and SNP-heritability for each of the five personality traits was calculated [29]. To investigate the relation among the different personality traits, the LDSC based correlation was also calculated between each pair of traits [30]. LDSC was also used to calculate genetic correlation of the personality traits with multiple other phenotypes (1437 traits) with CTG-VL webtool [31].

### Local genetic correlations

We used LAVA [15] to calculate local genetic heritability for the 5 personality traits and local genetic correlations for each pair. The genome was divided into 2,495 genomic chunks/loci to attain minimum linkage disequilibrium between them and maintain an approximate equal size of around 1 MB. The local genetic heritability of each of the 5 personality traits was calculated for each of the 2,495 loci. For a given personality trait pair, local genetic correlations were calculated only for pairs which had significant local genetic heritability (Bonferroni corrected P-value at 5% FDR) for both traits of the pair. Bonferroni multiple testing correction was also applied to genetic correlated P-value to consider significant correlated pairs.

### Transcriptome Wide Association Study

FUSION software [16] was used to perform TWAS. FUSION first estimates the SNP-heritability of steady-state gene and uses the nominally significant (P<0.05) genes for training the predictive models. The predictive model with significant out-of-sample *R*^2^ (>0.01) and nominal *P* < 0.05 in the 5-fold cross-validation was then used for the predictions in the GWAS data. The process is performed for all 5 personality EUR GWAS data with 14000 unique genes spanning over the 13 selected tissues. The expression weight panels for 13 a priori selected tissues were taken from GTEx v8 [17]. We selected the different available brain tissues and whole blood as the tissues of interest. where it Bonferroni corrections at FDR < 0.05 was applied with the 10,386 genes test for the 13 tissues to find the genes with significant hits (P<3.7X10^-9^).

### PWAS

We performed PWAS to test the association between genetically regulated protein expression and different personality traits individually using FUSION software [16]. The weights for genetic effect on protein expression for the PWAS were from Wingo *et al.* study [32]. In the PWAS, we integrated the protein weights with the summary statistics from the GWAS of each of the personality trait, respectively. Next, to decrease the probability of linkage contributing to the significant association in the PWAS, we performed colocalization analysis using COLOC [33]. In COLOC, we determined if the genetic variants that regulate protein expression colocalize with the GWAS variants for the personality trait. Significant proteins in the PWAS that also have COLOC PP4 >50% have a higher probability of being consistent with a causal role in the personality trait of interest.

### Fine mapping

To identify likely causal variants, we performed variant fine mapping using Polyfun software [19]. Since the fine-mapping was performed on the same EUR-ancestry data, SNP-specific prior causal probabilities were taken directly from the pre-computed causal probabilities of 19 million imputed UKB SNPs with MAF>0.01 based on 15 UK BioBank traits analysis. The fine mapping was performed on the GWAS sumstats for each of the 5 personality traits. SuSiE [34] was used to map the posterior causal probabilities of the SNPs. The SNPs with PIP value > 0.95 were considered as significant for neuroticism while a more relaxed cutoff of PIP > 0.80 was used for other 4 personality traits to avoid loss of causal variant information due to the relatively less power in their respective datasets.

### Conditional analysis

Conditional analysis was performed to investigate the possible confounding of depression or anxiety in neuroticism. Neuroticism meta data GWAS summary statistics was used and conditioned on MDD and anxiety in individual runs. The MDD summary statistics were from Levey et al. study [35] which includes a meta-analysis from the MVP, UKB, Psychiatric Genomics Consortium (PGC), and FinnGen. The Anxiety summary statistics were taken from Levey *et al.* study [24]. With depression/anxiety studies as covariate traits, the conditional analysis of neuroticism (target trait) was carried out using mtCOJO utility of GCTA [36]. Similarly, the same method was used to perform conditional analysis of agreeableness on MDDand anxiety.

### Drug Perturbation Analysis

FUMA was used to carry out the MAGMA based gene-association tests to find significantly associated genes for a trait from its GWAS summary data. Drugs were searched for both neuroticism and MDD individually using their respective significantly associated genes derived from neuroticism meta analysis summary statistics and MDD GWAS summary statistics from Levey et al. summary statistics. To predict drug candidates for a given trait, significant genes associated with neuroticism/depression ware given as input to gene2drug R-package [20]. Precomputed Pathway Expression Profiles of the Connectivity Map (Cmap) data were taken from DSEA website. For each query gene, maximum of 5 predicted drugs were predicted. Further, the drugs showing EScore > 0.5 and P-value less than 10^-6^ were considered significant. The process was repeated for MDD.

### Mendelian Randomization (MR)

MR was performed to study the causal relationship between 4 pairs of traits: neuroticism and MDD, neuroticism and anxiety, agreeableness and MDD, agreeableness and anxiety. These traits had the highest genetic correlation. The summary statistics described previously for conditional analysis for all four traits were used for carrying out MR analysis as well. TwoSample MR package was used to perform the MR [37]. For each pair of traits, the TwoSample MR was run twice to see the effect of exposure of each of the two traits on the outcome of other. After harmonizing the exposure and outcome instruments sets, clumping of SNPs (distance=500kb, r2=0.05) was performed before conducting the MR analysis. Because some of our samples included in the analysis of personality overlap with our outcomes and exposures of interest, and TwoSample MR is not robust to sample overlap, we also performed a sensitivity analysis for each trait pair using the MRlap package [21]. MRlap is specifically designed to account for many assumptions of MR, including sample overlap. It first calculates observed MR based effect values and then a corrected effect value by using the genetic co-variance calculated by ldsc.

## Discussion

We conducted a GWAS meta-analysis study of each of the “Big Five” personality traits in a sample size of up to 682,688 participants. We combined original GWAS results from the MVP (available for all five traits) with summary statistics from the UKB (neuroticism only) and GPC (all traits except neuroticism) cohorts to perform a well powered meta-analysis for EUR GWAS in each trait. We identified 468 independent significant SNPs associations mapping to 158 independent genomic loci of which one-third are novel. We identified 231 significant gene associations with neuroticism in the gene based analysis. The current study was also successful in identifying 23 significant genomic locus associations for the four other personality traits studied for which prior knowledge in the literature was very limited. For AFR we identified one GWS variant for agreeableness. This is likely a reflection of low power and underlines the critical need to increase recruitment in underrepresented groups. Our work provides new data to inform the underlying genetic architecture of personality traits.

Neuroticism, the trait with the largest available sample size in this study, is characterized by emotional instability, increased anxiousness, and low resilience to stressful events. As such, it has been the focus of previous efforts in GWAS studies. As seen previously, neuroticism overlaps substantially with psychopathology, where it is usually viewed as a precursor, or risk factor, for depressive and anxiety symptoms. Extraversion had the second largest sample size and had the highest SNP-based heritability in the MVP. In our data, scoring high on extraversion was genetically correlated with risk taking behaviors and had the second strongest negative genetic correlation with neuroticism. Agreeableness assays how someone relates with other people, ie how trusting one is or how likely to find fault in others. This trait was the most negatively correlated with neuroticism and irritability as well as Major Depressive Disorder, anxiety, and manic symptoms. Conscientiousness items relate to discipline and thoroughness, with specific questions being ‘are you lazy’ and ‘does a thorough job’. This trait was most closely associated with ‘types of physical activity in last 4 weeks: Heavy DIY (do it yourself).’ Finally, openness BFI-10 items assay imagination and artistic interest. Openness was positively associated with extraversion and risk taking in our data. Educational attainment was positively correlated with openness and negatively associated with neuroticism, while the other 3 personality traits showed essentially no such overlap (Figure 5). Since these are self-report items, they naturally reflect one’s own assessment of one’s personality traits, which might filter actual traits and behavior through a lens of how one wishes to appear or be perceived.

**Figure 5:**
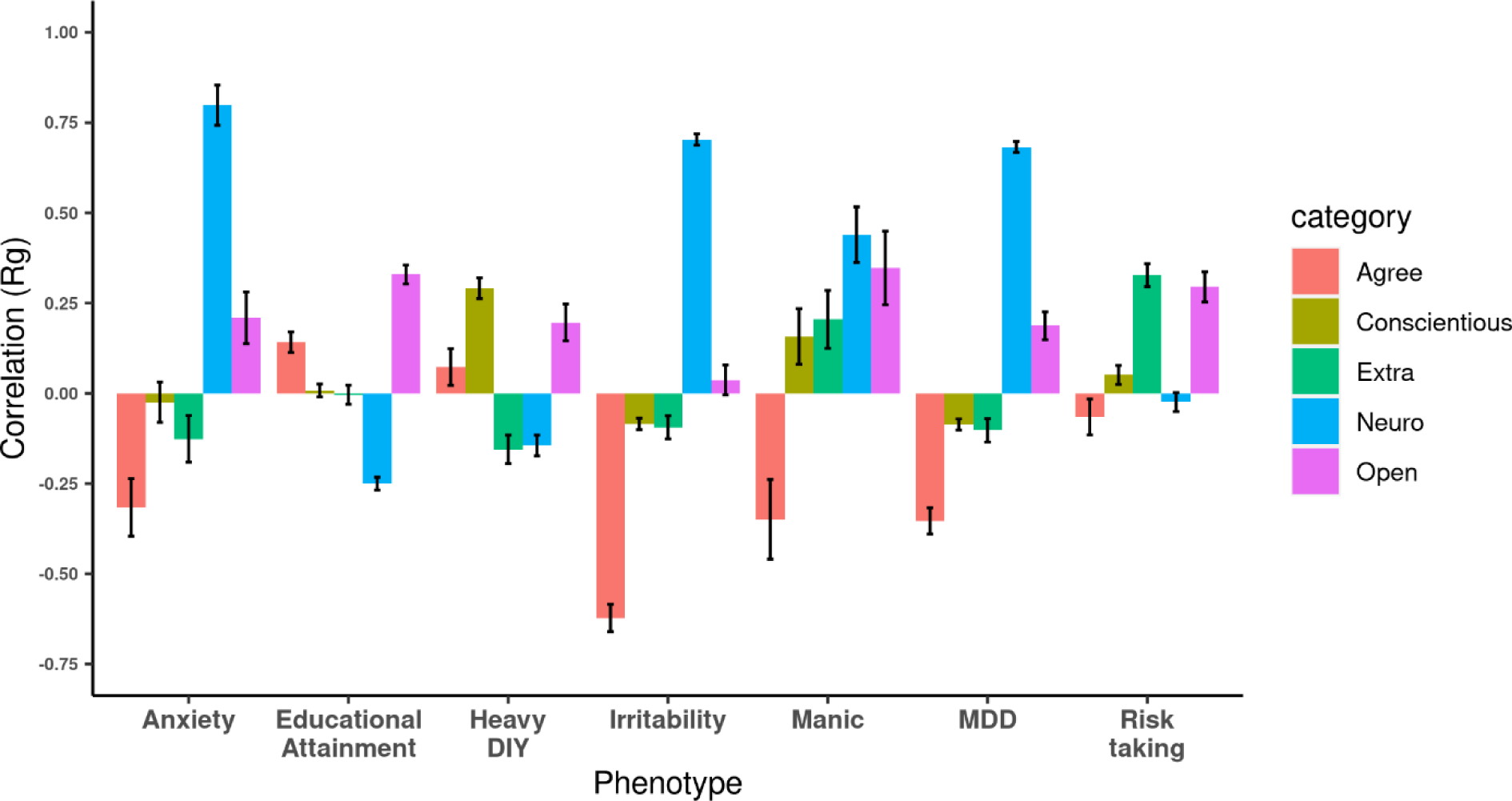
Bar chart with genetic correlation of each of the five personality trait with different psychological disorder/trait or different behaviors plotted. Anxiety: Substances taken for anxiety; Medication prescribed to you (for at least two weeks). Heavy DIY: Types of physical activity in last 4 weeks: Heavy DIY (eg: weeding;lawn mowing;carpentry;digging). Manic Behaviour: Ever Manic/hyper behaviour for 2days. MDD: Major depressive disorder.

Using these GWAS summary statistics, with excellent power for neuroticism and moderate power for the other traits, we investigated the genetic heritability of the different personality traits and studied genetic correlations among them using LDSC. SNP-based heritability for all 5 personality traits in EUR were statistically significant. Out of all the personality pairs studied the strongest relationship was a negative genetic correlation observed between neuroticism and agreeableness (rG=-0.51, Figure 1B). Examining the genetic correlations of the 5 personality traits with 1437 external traits including depression (Neuroticism rG=0.68, Agreeableness rG=- 0.35), manic behavior (Neuroticism rG=0.44, Agreeableness rG=-0.35), anxiety (Neuroticism rG=080, Agreeableness rG=-0.33), and irritability (Neuroticism rG=0.70, Agreeableness rG=- 0.62) further reflected a pattern of opposing relationships between these traits (see Figure 5 and Supplementary Sheet 3). We also calculated local genetic correlations between personality pairs using LAVA, which helped in identifying the genomic regions playing roles in differential overlap in the genetic architecture of personality. This analysis identified several regions where the effect direction differs from the whole genome genetic correlation.

The MVP, our discovery dataset, is one of the world’s largest biobanks and is a valuable resource for genetic studies. Some previously published personality trait studies have significant contribution from UKBiobank data. It is important to quantify the heterogeneity in these independent cohorts and the different definitions of personality phenotype within each. We investigated the genetic correlation between traits defined based on different inventories (BFI-10, EPQ-RS, NEO-FFI) of personality ascertainment with different cohorts-MVP, UKBiobank, GPC, respectively. For neuroticism, UKB and MVP studies showed a high rG value of 0.80 making these two independent cohorts suitable for meta-analysis (see Table S1). Similarly, for extraversion, NEO-FFI and two item inventories showed high rG of 0.89 in the extraversion data of GPC and MVP studies. While for agreeableness, openness, and consciousness, the rGs between MVP and GPC cohort was lower (0.63-0.72); this may be due to the small size of the GPC dataset for these traits. No novel loci were identified in the meta-analysis with GPC for these traits.

TWAS revealed common genes with changes in gene expression but with opposite direction of effect for some personality traits. A study by Ward *et al.* in 2020, reported 5 of these genes as eQTLs showing significant associations with mood instability [38]. This is further supported by the local genetic correlation studies (see Suppl. Sheet S3) where we found genomic region 45883902-47516224 on chromosome 17, which harbors genes *KANSL1-AS1, MAPT, MAPT-IT1*, showing negative local genetic correlation between neuroticism and extraversion with a rho value of -0.57 and r^2^ value of 0.32. rs1876829, which maps to CRHR-Intronic Transcript 1, emerged as the lead SNP (P=7.872e-39) for neuroticism in the GWAS analysis. We also found multiple eQTL SNPs in this genomic region (rs8072451, rs17689471, rs173365, rs11012) for the CRHR1 gene to be significantly associated (P value ranging from 10^-5^ to 10^-37^). The TWAS analysis showed significant association of this gene with neuroticism in nervous system tissues including caudate basal ganglia, frontal cortex, hippocampus, spinal cord cervical region. *CRHR1* encodes the receptor of corticotropin releasing hormone family which are major regulators of the hypothalamic-pituitary-adrenal pathway [39]. Genetic variation in the CRH system has been linked to several psychiatric illnesses [40]. Another study reported hypermethylation at CRH-associated CpG site-cg19035496 in individuals with high general psychiatric risk score for disorders like depression, anxiety, post-traumatic stress disorder, obsessive compulsive disorder [41]. Further, study by Gelernter *et al*. found *CRHR1* significantly associated with re-experiencing PTSD symptoms [42] and also maximum habitual alcohol intake [43]. This gene is also involved in hippocampal neurogenesis [40] while reduced hippocampal activation is associated with elevated neuroticism [44]. This makes *CRHR1* a good lead candidate to be followed in future studies to understand the molecular processes impacted by genetic variation underlying a range pf psychiatric traits including neuroticism.

While gene expression associations give a wide array of information on the involvement of different genes regulating the different biological processes underlying the biology of traits, searching protein expression associations confers several advantages as proteins are the final executioners in the functioning of all cells for many biological processes. Through PWAS studies, we found 47 proteins showing significant association with neuroticism in the dlPFC. The PWAS analysis also identified Leucine-rich repeat and fibronectin type III domain containing 5 (LRFN5) protein’s association with neuroticism and this protein is also involved in synapse formation. This protein has shown higher levels in MDD patients and has been suggested as a potential MDD biomarker [45].

Examples of genes for which we found converging evidence in neuroticism for transcript and protein level associations with neuroticism include low density lipoprotein receptor-related protein 4 (*LRP4),* syntaxin 4 (*STX4*), and metabolism of cobalamin associated B (*MMAB*) (Supplementary Sheet 8). *LRP4* has diverse roles in neuromuscular junction and in disorders of the nervous system including Alzheimer’s disease and amyotrophic lateral sclerosis [46], *STX4* is implicated in synaptic growth and plasticity [47], and *MMAB*, which catalyzes the final step in the conversion of cobalamin (Vitamin B12) into adenosylcobalamin (biologically active coenzyme B12), all of which have broad implications for brain function including those in relation to methylmalonic acidemia [48]. Low levels of plasma vitamin B12 have been found to be associated with higher depression cases in multiple studies [49].

We investigated the relationship of these personality traits with other psychiatric traits, cognitive functions and disorders in a broad phenome-wide scan of genetic correlations with 1,435 traits. 325 traits showed significant genetic correlations with at least one of the five personality traits following multiple testing correction. Two important traits that had some of the strongest associations were MDD and anxiety. Whereas the association of neuroticism with depression and anxiety has been previously considered [4, 11], our analysis revealed that another personality trait -agreeableness – is also strongly associated with both anxiety and depression but in the opposite direction to neuroticism, showing a potential protective relationship. Mendelian randomization indicated a strong bidirectional causal relationship between neuroticism with anxiety and depression while showing a bidirectional protective relationship for agreeableness for both traits. Variance explained for neuroticism was attenuated upon conditioning for MDD but remained significant, indicating some independent genetic component for neuroticism despite the strong overlap. Similar, but less strong effect was seen of anxiety on neuroticism, which may be partly due to lower power of available anxiety summary statistics. Larger studies of anxiety disorders are needed to better understand this relationship. Conversely, when we conditioned on agreeableness, for MDD and anxiety we did not observe a significant change in SNP-based heritability We conducted mendelian randomization to further discern these patterns and it showed bidirectional causal effects with neuroticism, confirming a high degree of inter-relatedness between the traits. Given the high degree of genetic overlap between trait neuroticism and the expectation of personality trait expression preceding age of onset for MDD, a high trait neuroticism may be considered an early risk factor for anxiety, depressive and related psychopathology. Indeed, studies have shown persistent elevated neuroticism through adolescence is a risk factor for later susceptibility to anxiety and MDD diagnosis [50].

Personality traits are known to have complex interactions with other human behaviors. In this work we have conducted comprehensive genomic studies of personality traits. We performed a genome-wide association study in the MVP sample, the largest and most diverse biobank in the world, in both European and African ancestry to better understand genetic factors underlying personality traits. We combined this information with previously published results in a large meta-analysis, identifying novel genetic associations with five personality traits studied. We identified interactions in a phenome-wide genetic correlation analysis, finding novel relationships between complex traits. We used in-silico analysis techniques to identify genetic overlap and causal relationships with depression and anxiety disorders. We also characterized underlying biology using predicted changes in gene and protein expression, biological pathway enrichment, and drug perturbation analysis. These results substantially enhance our knowledge of the genetic basis of personality traits and their relationship to psychopathology.

## Supporting information

Suppl. sheet 1 AFR Ancestry

Suppl. sheet 2 Meta-analysis genomic loci

Suppl. sheet 3 Personality-traits correlations

Suppl. sheet 4 local genetic correlations

Suppl. sheet 5 TWAS

Suppl. sheet 6 PWAS

Suppl. sheet 7 Finemapping

Suppl. sheet 8 Drug response

Suppl. sheet 9 TWAS-PWAS convergence

## Data Availability

All data produced in the present work are contained in the manuscript

## Acknowledgements

This research is based on data from the Million Veteran Program, Office of Research and Development, Veterans Health Administration, and was supported by award #5IK2BX005058. This publication does not represent the views of the Department of Veteran Affairs or the United States Government.

AW was supported by a BLRD CDT Award from the US Department of Veterans Affairs # 1IK4BX005219.

AW and TW were supported by R01 grant # AG072120.

Detailed MVP Core team acknowledgements are included in the supplement.

